# Comparison of the audiological knowledge of three chatbots – ChatGPT, Bing Chat, and Bard

**DOI:** 10.1101/2023.11.22.23298893

**Authors:** W. Wiktor Jedrzejczak, Krzysztof Kochanek

## Abstract

**Objective:** The purpose of this study was to evaluate three chatbots – OpenAI ChatGPT, Microsoft Bing Chat, and Google Bard – in terms of their responses to a defined set of audiological questions.

**Design:** Each chatbot was presented with the same 10 questions. The authors rated the responses on a Likert scale ranging from 1 to 5. Additional features, such as the number of inaccuracies or errors and the provision of references, were also examined.

**Results:** Most responses given by all three chatbots were rated as satisfactory or better. However all chatbots generated at least a few errors or inaccuracies. ChatGPT achieved the highest overall score, while Bard was the worst. Bard was also the only chatbot unable to provide a response to one of the questions. ChatGPT was the only chatbot that did not provide information about its sources.

**Conclusions:** Chatbots are an intriguing tool that can be used to access basic information in a specialized area like audiology. Nevertheless, one needs to be careful, as correct information is not infrequently mixed in with errors that are hard to pick up unless the user is well versed in the field.

## 1. Introduction

Although the term artificial intelligence (AI) and certain applications of it have been known for many years [1], tools based on it have recently become very popular. A major trigger was the introduction in 2022 of ChatGPT, which was soon made publicly accessible by its developers, OpenAI. ChatGPT stands for Chat Generative Pre-trained Transformer. ChatGPT is a so-called ‘chatbot’ – a conversational, natural language processing (NLP) system of AI based on large language models (LLMs) [2,3]. The intent of the chatbot is to interact with users in a way that resembles a conversation between humans. ChatGPT has generated a lot of interest – 100 million users in 2 months [4]. This has prompted the release of other chatbots such as Google Bard and Microsoft Bing Chat.

ChatGPT, Bing Chat, and Bard share numerous capabilities and can be utilised in similar ways. They are based on the transformer-type model architecture, a concept that has recently revolutionized the AI field [5]. This architecture incorporates a self-attention mechanism, enabling the model to focus on various parts of the input sequence with varying levels of attention. These models are pretrained on extensive datasets and fine-tuned before being made widely available. However, ChatGPT, Bing Chat, and Bard employ different models. ChatGPT utilizes the GPT-3.5 model, trained on information sources up to 2021. Bing Chat uses the GPT-4 model, which is more advanced than GPT-3.5 but belongs to the same line of models developed by OpenAI. Bard employs the PaLM 2 model, developed by Google (PaLM stands for Pathways Language Model). Unlike ChatGPT, both Bing Chat and Bard have access to live data on the Internet. All three systems are still in development and may not be permanently or temporarily available in all countries due to various reasons, such as governmental decisions or software provider policies. For more detailed information, readers are encouraged to review Motlagh et al.’s comprehensive comparison of different chatbots [6]. While there are numerous other chatbots and systems based on similar principles for various purposes (such as graphics), this discussion focuses solely on the aforementioned three.

Enthusiasm for chatbots continues to grow, and their potential in various applications is under discussion. For example, ChatGPT recently achieved a score of 60% on the United States Medical Licensing Examination, giving it a clear pass [7]. Some recent press reports have tried to link the rise in ChatGPT use with the start of the school year [8]. There are cases of researchers publishing texts generated by chatbots as their own [9]. Software has been created with the purpose of identifying not just plagiarism but also text generated by AI [10]. Scientific journals have begun to ask for a declaration that AI was not used in the preparation of a submitted manuscript. So far, the only allowed form of AI use is for correcting text, but in such cases it must be acknowledged.

Regardless of the controversy, the fact is that people are already using chatbots. So aside from discussion about the usefulness, dangers, rules, and ethical boundaries, it would be useful to evaluate the performance of chatbots on various tasks. Unfortunately, most papers just seem to discuss the problem. There appear to be only a relatively small group of studies that have endeavored to evaluate the actual performance of chatbots or compare their answers. For example, when searching PubMed with the query (chatgpt[Title]) we received 1108 hits for the year 2023 (search made 06.10.23). However, when searching for (chatgpt[Title]) AND ((performance[Title]) OR (evaluation[Title]) OR (comparison[Title])) we received just 102 hits. This rapid comparison shows the large imbalance between papers that actually try to evaluate chatbots with others that just present personal reviews or opinions. It is our perspective that scientists ought to cease discussion on the matter and engage in substantive empirical testing.

Confining our attention to educational uses, a quick review made 3 months after the release of ChatGPT [11] indicated that its performance depends on the particular topic. There are reports of outstanding performance in areas such as economics [12] and programming [13], whereas its results in mathematics are typically unsatisfactory [14]. In the case of medicine, there are reports of chatbots achieving good results for questions related to cardiology [15] and psychiatry [16].

In audiology, chatbots could potentially be used by patients, students, clinicians and researchers [17]. Swanepoel and colleagues identified possible applications as well as risks, and gave some examples by different groups of users. For example, they suggest that audiological patients could use chatbots for initial screening and recommendation of interventions, education and support, reminders and follow-up communications, or teleaudiology services. Presently, there seem to be no other literature on chatbots in audiology although there are already some in the closely related field of otolargyngology. For example, there is one study investigating the usefulness of ChatGPT as a source of patient information [18], and another that investigated the possibility, for educational purposes, of using ChatGPT to generate examples of dialogues between patient and otolaryngologist [19].

In summary, despite many discussions there is no consensus about the usefulness of chatbots. Although they may be quite good in conversation and in certain general topics, it is not clear how they perform in fields like audiology, which is not as broad as other medical fields. For example, when checking the Journal Citation Reports (a tool for the assessment of scholarly journals provided by Clarivate) in the group of Clinical Medicine journals, there are only 27 journals belonging to the category of Audiology & Speech-Language Pathology, while for the category of Psychiatry, there are 156, and for the category of Cardiac & Cardiovascular Systems, there are 143 (in the current year of 2023). In this way, each of these two mentioned fields has an output approximately five times larger than Audiology & Speech-Language Pathology. Since chatbots’ knowledge is based on available resources, it is likely that their capabilities in a field such as audiology may differ from those in broader fields, as seen in previous studies [15,16]. To complicate the issue, there are some reports indicating that, for certain purposes, some chatbots are better than others [20]. So far as we are aware, there are no studies of how chatbots perform in response to audiological questions. This study attempts to fill this gap and contribute valuable insights to the broader discourse on the efficacy of chatbots in specialized fields.

The purpose of this study was to ask ChatGPT, BingChat, and Bard the same set of questions from a variety of topics in audiology and compare their answers. It is hoped that such an approach will aid in understanding the limitations of chatbots and identifying areas for improvement.

## 2. Materials and Methods

The responses of three chatbots to a set of questions were evaluated. The chatbots were OpenAI ChatGPT (version 3.5), Microsoft Bing Chat (which is based on ChatGPT but with some modifications), and Google Bard. Standard publicly available versions were used. Bard was marked as an experimental version, and there were some problems with access in our country (Poland). A total of 10 questions (Table 1) were prepared, roughly categorized as basic (questions 1–3), intermediate (4– 6), and specialised (7–10). The questions were deliberately diverse, being either short and general or long and forcing a more detailed answer. Questions in the basic and intermediate groups are those a patient or student might ask, while specialized questions are those that a physician, researcher, or hearing specialist might seek answers to. The questions were presented to the three AI models on 28.08.2023.

**Table 1.** Questions used to test chatbots.

| No | Question | Type |
| --- | --- | --- |
| 1 | What is an audiometric hearing test? | Basic |
| 2 | What is cochlear implant surgery? |  |
| 3 | What is newborn hearing screening? |  |
| 4 | What is air-bone gap? | Intermediate |
| 5 | What is auditory neuropathy and how is it diagnosed? |  |
| 6 | Does the stapedius reflex protect the inner ear from damage? |  |
| 7 | What are the clinical signs of conductive hearing loss? | Specialized |
| 8 | Does it make sense to do a auditory brainstem responses examination if an MRI confirmed large cerebello pontine angle tumor in one ear? |  |
| 9 | What is the therapeutic management of acoustic neuroma? |  |
| 10 | What is hidden hearing loss? |  |

The correctness of the questions was then evaluated by two experts (the authors) using a 5-point Likert scale (1 = extremely unsatisfactory, 2 = unsatisfactory, 3 = neutral, 4 = satisfactory, and 5 = extremely satisfactory). The experts went over all the responses twice and agreed on the scores. The answers were evaluated in a general way, according to correctness and completeness, the same as in a situation where a student was answering the question. The similar evaluation method was employed in several previous studies on chatbot performance, e.g. [21,22,23]. Other features were also evaluated like number of errors, inaccuracies, whether information sources were provided, whether a suggestion was made to refer to a specialist, and the number of words (counted using Microsoft Word). We decided to rate small mistakes as inaccuracies when the response was imprecise although not misleading. The responses of the chatbots are provided in Supplementary Files 1, 2, and 3. As well as the actual chatbot responses, we also provide scores, errors, and number of inaccuracies.

All analyses were made in Matlab (version 2020a, MathWorks, Natick, MA). All datasets were tested for normality of distribution by a Shapiro–Wilk test. If the tests were passed, a t-test was then used, otherwise a nonparametric Mann–Whitney U-test. In addition, an ANOVA or its nonparametric equivalent Friedman test was used. In all analyses, a 95% confidence level (p < 0.05) was taken as the criterion of significance.

## 3. Results

The results of evaluating the chatbots are presented in Table 2, which shows average results as well as the results of statistical comparisons. All individual scores and other ratings are in supplementary file 4.

**Table 2.** Results of evaluation of the chatbots. The last column shows the result of statistical test if there was a difference between three chatbots. STD – standard deviation.

|  | ChatGPT | BingChat | Bard | *p* value |
| --- | --- | --- | --- | --- |
| Score (min = 10, max = 50) | 45 | 37 | 36 | 0.10 |
| Number of responses rated as 5 (max = 10) | 6 | 1 | 3 | 0.057 |
| Number of responses rated as 4 or better (max = 10) | 9 | 6 | 6 | 0.24 |
| Number of errors | 2 | 1 | 6 | 0.81 |
| Number of inaccuracies | 4 | 4 | 2 | 0.58 |
| Number of errors and inaccuracies together | 6 | 5 | 8 | 0.94 |
| Number of responses in which sources were provided (max = 10) | 0 | 8 | 2 | 0.0004 |
| Number of responses in which the help of a specialist was suggested (max = 10) | 4 | 1 | 9 | 0.0014 |
| Number of words (STD) | 472 (49) | 112 (46) | 356 (45) | 0.0001 |

Most of the responses given by all three chatbots were rated as satisfactory or better. However all chatbots generated at least a few errors or inaccuracies. ChatGPT achieved the highest overall score, while Bard was the worst. Bard was also the only chatbot unable to provide a response to one of the questions. Nevertheless, there were no statistically significant differences between scores (even when they were divided against types – easy/intermediate/specialized), numbers of errors, and number of inaccuracies.

The scores each chatbot received were compared to a mid-rating of 3 (i.e. a total score of 30 for 10 responses). Our expectation was that a competent answer set should achieve a better rating than that. ChatGPT and Bing were significantly better than 3 (p = 0.0002 and 0.0054 respectively), while Bard was not significantly better (p = 0.094).

It should be noted that Bard was the only chatbot which always suggested that a specialist should be consulted. Also notable is that ChatGPT was the only chatbot that did not provide any information about sources. Bard provided sources for 2 of its 10 responses while Bing gave them for 8. Furthermore, Bing, besides providing a source, also usually provided several additional links to pages where knowledge on a topic could be deepened.

There were significant differences between chatbots in the length of responses. Despite the fact that the questions had diverse topics and structures, each chatbot always provided responses of similar length. Furthermore the variability in length of each chatbot’s answers was similar: even though the numbers of words differed significantly, the standard deviations were much the same. ChatGPT gave the longest answers while Bing gave the shortest.

It is revealing to examine examples of specific errors made by each chatbot. In most cases the mistakes were hidden among well-constructed text, which was in large measure correct. Here, we present some excerpts from responses, which in some cases were quite long. For the full responses, the reader is referred to supplementary files 1, 2, and 3.

As a first example, in response to question 8, ChatGPT said: “Performing an ABR test in the presence of a large CPA tumor may carry certain risks, such as discomfort or potential complications related to the testing procedure. It is essential to weigh these potential risks against the expected benefits, especially when the primary goal is to address the tumor itself.” This is untrue, since auditory brainstem response (ABR) testing is known to be a very safe noninvasive method without any side effects. ChatGPT did not say what the complications might be. Such an answer could be potentially harmful if a patient were to decline a test fearing some imagined complication.

In response to question 3, Bing said that “The newborn hearing test is called the automated otoacoustic emission (AOAE) test.” While otoacoustic emissions (OAEs) are in-deed often used in hearing screening they are not synonymous with ‘newborn hearing test’ as other tests may be also performed, such as ABRs.

In response to question 1, Bard said that “A normal audiogram shows a smooth, sloping line.” This is an error, as a normal audiogram is usually a straight line (and importantly it is above 20 dB HL), whereas a sloping line indicates age- or noise-related hearing loss.

Interestingly, in response to question 6, which can be answered simply with “yes” or “no,” none of the chatbots provided such a brief response. Instead, each of the chatbots offered some explanation, even though they were not explicitly asked to do so.

There were also minor mistakes we called inaccuracies. For example, in response to question 1 all chatbots said that hearing thresholds were evaluated in decibels (dB) but they did not say that these are actually dB hearing level (dB HL). By itself this statement is not erroneous but a more precise answer would say dB HL.

## 4. Discussion

When looking at chatbot responses to audiology questions one’s first impression might be positive. All the chatbots provided nicely framed responses. However closer inspection shows some serious problems.

It is not easy to evaluate all three chatbots using the same grading system. For example, ChatGPT gives long answers with considerable detail, while Bing Chat provides short, almost telegraphic responses. Bard falls somewhere in the middle. Bing’s short responses are not necessarily a bad thing, as there is no rule as to how full a response needs to be. Therefore we evaluated a response positively if it provided substantial content, regardless of length, but did not contain errors. In general, we rated the majority of responses given by the chatbots as satisfactory. Since the scores we assigned are subjective, perhaps the number of errors and inaccuracies might be a more objective measure of chatbot performance. We think it is worth underlining that, despite the overall positive score, all chatbots generated some errors or inaccuracies. Such limitations might have different consequences depending on who is searching for the information. Someone with familiarity with the topic might readily detect mistakes, but someone new to the topic might be misled. Interestingly, we did not notice large differences in responses in terms of the difficulty of the questions. Errors occurred in both simple and specialized questions. It is possible that such a difference would be more noticeable if there were more questions of each type.

Looking more closely at each of the chatbots, each has unique characteristics. For example, whereas the average number of words in responses differed, the standard deviations were similar. This means that each chatbot generated responses according to a predetermined pattern using more or less the same number of words. There also seemed to be some standard word patterns [24,25]. Perhaps after further use it might be possible to recognize which response came from which chatbot. ChatGPT provided the longest responses, often giving additional background. However, sometimes it strayed off topic. The major limitation of ChatGPT is that it does not provide references. For some questions (but not the case for our standard set) ChatGPT answered that “my knowledge is not current beyond September 2021”. In our opinion it is good for a system to know its own limitations. Bing Chat provided the shortest responses, which were sometimes sketchy; however they were usually free of errors and to the point. The greatest advantage of Bing is that it provides references and suggestions for additional sources of information. Bard responses were longer and were more similar to those given by ChatGPT, and they sometimes contained references. However, Bard was the only one to not answer a question. In response to question number 3, it said: “I’m unable to help you with that, as I’m only a language model and don’t have the necessary information or abilities.” At the same time, Bard was the only chatbot to suggest referring to a specialist in all the other responses it provided. We regard this as important, as it reminds the user that chatbot is only a tool with limited capabilities, and the disclaimer is some sort of protection against errors. Note also that Bard is marked as an experimental version so it is possible that in future the final product will perform better. Our experience with chatbots can be summarized with the suggestion to use Bing Chat if searching for a short answer and to use ChatGPT if wanting more comprehensive information.

When thinking of possible applications of chatbots in audiology it seems hard to point to a likely user right now. In the future, when the performance of chatbots has improved, more possibilities will no doubt appear, as recognized by Swanepoel et al. [17]. Currently, any application to science or education is ruled out due to a lack of reference to sources, although it is true that in normal conversation one does not usually cite sources. In science, however, providing a reference for a statement is crucial. This lack seems to be the current main limitation. In our tests, only one of the responses contained a reference to a research paper. Only Bing Chat provided sources for its responses, which were supplied in most cases. However, the sources were typically links to webpages (usually unverified information) not scientific papers (verified information). On the positive side, Bing Chat’s suggested webpages often provided references to scientific papers or books.

Although questions were asked in English, we also tested asking some in our native Polish language. Surprisingly, the responses differed considerably. Bing Chat appeared to use different sources (giving Polish language webpages), although there were many more errors and inaccuracies. Based on our experience we recommend using English for asking scientific questions and then translating the answers if required (possibly using Google Translate or DeepL).

Some studies have already compared the performance of different chatbots. For example, Patil et al. [22] compared ChatGPT and Bard for radiology questions and found that ChatGPT gave better results. In hematology, ChatGPT achieved the highest score, followed by Bard and then Bing [26]. Also ChatGPT responses was rated higher than both Bard and Bing when solving case vignettes in physiology [27]. On the other hand, when predicting drug interactions, Bing outperformed Bard and ChatGPT [28]. When providing information on rhinoplasty, Bard excelled over ChatGPT and Bing [23]. In our present study, ChatGPT was rated highest although the number of mistakes was similar across the three chatbots. It could be that different chatbots may perform better in distinct areas, but taken together the rankings do not seem to be clear-cut.

The main limitation of this study is that we subjectively rated the responses. We tried to be as objective as possible but some subjectivity is inevitable. We applied our best knowledge but it is possible we made errors or overlooked something. All the chatbot responses are attached as supplementary files so that the reader can make their own assessment. Such an evaluation is similar to how student or research papers are evaluated. A second limitation is the choice of questions. Obviously, responses to a different set of questions would generate different results. As said earlier, we deliberately tried to create questions on various topics, and of different difficulty and length, in order for the test to be as broad-based as possible.

In our work, we simply compared responses to certain open questions. There are other ways of testing, including multiple choice or finding errors in lengthy texts. We did not develop extended conversations, or add supplementary questions to delve deeper into a topic. It is possible that evaluating the responses of a chatbot to multiple questions on a single topic would yield different scores. The difficulty is that the number of questions needed to fully cover a topic is likely to vary between chatbots, so comparisons would be hard to make equal. This is the reason we kept to a single question.

When chatbots are considered for educational or scientific uses, we are thinking of it being used as a source of knowledge or as a test of certain scenarios. We strongly advocate against their unethical usage (e.g. to pass an exam or ghost write a scientific paper). Unfortunately such uses have been already documented, where papers have been found to have the phrase ‘regenerate response’ from the interface of ChatGPT [29,30]. The international community is currently engaged in a discussion about the boundaries of chatbot use. Some voices are advocating for the restriction of chatbot usage, and there are even calls to halt further AI development altogether. The arguments against it include ethical concerns, security risks, lack of regulation, and the potential loss of human control.

Ultimately, the responsibility lies with us to determine how technology is employed, whether for positive or negative purposes.

## 5. Conclusions

Each tested chatbot had its positives and negatives, and it is hard to say which is the best. Bing Chat seems best when searching for short answers while ChatGPT is better when searching for more comprehensive information. Bard was rated the lowest, but it may be because it is currently an experimental version.

Chatbots were created for conversation and are not necessarily suited to scientific uses. Currently, we should recognize the role of chatbots in audiology as being more of a sort of initial information source, or as a tool to examine scenarios. Given the present limitations, is there anyone who could effectively use a chatbot for audiological purposes? Possibly, a student might want to ask a question and then check it over for accuracy. Or a researcher may converse with a chatbot to come up with new ideas. At the same time, however, for students or patients chatbots present many risks. Although the response may seem to be correct, if one is not an expert you can be easily misled, and the lack of references is a major obstacle to verification. We believe that chatbots are not there to create statements of fact, or to evaluate those of others. If they did, we would not be needed.

## Data Availability

All data produced in the present study are available as supplementary files.

## References

1. Haenlein, M., & Kaplan, A. (2019). A brief history of artificial intelligence: On the past, present, and future of artificial intelligence. California management review, 61(4), 5–14. 10.1177/0008125619864925

2. Adamopoulou, E., & Moussiades, L. (2020). Chatbots: History, technology, and applications. Machine Learning with Applications, 2, 100006. 10.1016/j.mlwa.2020.100006

3. Jeon, J., & Lee, S. (2023). Large language models in education: A focus on the complementary relationship between human teachers and ChatGPT. Education and Information Technologies, 1–20. 10.1007/s10639-023-11834-1

4. Trust, T., Whalen, J., & Mouza, C. (2023). ChatGPT: Challenges, Opportunities, and Implications for Teacher Education. Contemporary Issues in Technology and Teacher Education, 23(1), 1–23.

5. Vaswani, A., Shazeer, N., Parmar, N., Uszkoreit, J., Jones, L., Gomez, A. N., & Polosukhin, I. (2017). Attention is all you need. Advances in Neural Information Processing Systems, 30.

6. Motlagh, N. Y., Khajavi, M., Sharifi, A., & Ahmadi, M. (2023). The impact of artificial intelligence on the evolution of digital education: a comparative study of OpenAI text generation tools including ChatGPT, Bing Chat, Bard, and Ernie. arXiv preprint arXiv:2309.02029.

7. Gilson, A., Safranek, C. W., Huang, T., Socrates, V., Chi, L., Taylor, R. A., & Chartash, D. (2023). How does ChatGPT perform on the United States medical licensing examination? The implications of large language models for medical education and knowledge assessment. JMIR Medical Education, 9(1), e45312. doi: 10.2196/45312.

8. Bloomberg https://www.bloomberg.com/news/articles/2023-09-20/chatgpt-usage-is-rising-again-as-students-return-to-school?srnd=premium&leadSource=uverify%20wall (accessed on 11.10.2023)

9. Conroy, G. (2023). Scientific sleuths spot dishonest ChatGPT use in papers. Nature. 10.1038/d41586-023-02477-w

10. Rahimi, F., & Abadi, A. T. B. (2023). ChatGPT and publication ethics. Archives of medical research, 54(3), 272–274. doi: 10.1016/j.arcmed.2023.03.004.

11. Lo, C. K. (2023). What is the impact of ChatGPT on education? A rapid review of the literature. Education Sciences, 13(4), 410. 10.3390/educsci13040410

12. Geerling, W., Mateer, G. D., Wooten, J., & Damodaran, N. (2023). Is ChatGPT Smarter than a Student in Principles of Economics? Available at SSRN 4356034.

13. Buchberger, B. (2023). Is ChatGPT smarter than master’s applicants?: RISC Report Series, 23–04.

14. Frieder, S., Pinchetti, L., Griffiths, R. R., Salvatori, T., Lukasiewicz, T., Petersen, P. C., & Berner, J. (2023). Mathematical capabilities of chatgpt. arXiv preprint arXiv:2301.13867.

15. Skalidis, I., Cagnina, A., Luangphiphat, W., Mahendiran, T., Muller, O., Abbe, E., & Fournier, S. (2023). ChatGPT takes on the European Exam in Core Cardiology: an artificial intelligence success story?. European Heart Journal-Digital Health, 4(3), 279-281. doi: 10.1093/ehjdh/ztad029. Erratum in: Eur Heart J Digit Health. 2023 May 17;4(4):357.

16. Luykx, J. J., Gerritse, F., Habets, P. C., & Vinkers, C. H. (2023). The performance of ChatGPT in generating answers to clinical questions in psychiatry: a two-layer assessment. World psychiatry, 22(3), 479. doi: 10.1002/wps.21145.

17. Swanepoel, D. W., Manchaiah, V., & Wasmann, J. W. A. (2023). The rise of AI Chatbots in hearing health care. The Hearing Journal, 76(04), 26–30. DOI: 10.1097/01.HJ.0000927336.03567.3e

18. Nielsen, J. P., von Buchwald, C., & Grønhøj, C. (2023). Validity of the large language model ChatGPT (GPT4) as a patient information source in otolaryngology by a variety of doctors in a tertiary otorhinolaryngology department. Acta Oto-Laryngologica, 1–4. doi: 10.1080/00016489.2023.2254809.

19. Topsakal, O., Akinci, T. C., & Celikoyar, M. (2023). Evaluating Patient and Otolaryngologist Dialogues Generated by ChatGPT, Are They Adequate? 10.21203/rs.3.rs-2719379/v1

20. Dao, X. Q., & Le, N. B. (2023). Chatgpt is good but bing chat is better for vietnamese students. arXiv preprint arXiv:2307.08272.

21. Deiana, G., Dettori, M., Arghittu, A., Azara, A., Gabutti, G., & Castiglia, P. (2023). Artificial Intelligence and Public Health: Evaluating ChatGPT Responses to Vaccination Myths and Misconceptions. Vaccines, 11(7), 1217.

22. Patil, N. S., Huang, R. S., van der Pol, C. B., & Larocque, N. (2023). Comparative performance of ChatGPT and bard in a text-based radiology knowledge assessment. Canadian Association of Radiologists Journal, 08465371231193716. doi: 10.1177/08465371231193716.

23. Seth, I., Lim, B., Xie, Y., Cevik, J., Rozen, W. M., Ross, R. J., & Lee, M. (2023, September). Comparing the Efficacy of Large Language Models ChatGPT, Bard, and Bing AI in Providing Information on Rhinoplasty: An Observational Study. In Aesthetic Surgery Journal Open Forum (p. ojad084). Oxford University Press. doi: 10.1093/asjof/ojad084.

24. AlAfnan, M. A., & MohdZuki, S. F. (2023). Do artificial intelligence chatbots have a writing style? An investigation into the stylistic features of ChatGPT-4. Journal of Artificial intelligence and technology, 3(3), 85–94.

25. Guo, B., Zhang, X., Wang, Z., Jiang, M., Nie, J., Ding, Y., & Wu, Y. (2023). How close is chatgpt to human experts? comparison corpus, evaluation, and detection. arXiv preprint arXiv:2301.07597.

26. Kumari, A., Kumari, A., Singh, A., Singh, S. K., Juhi, A., Dhanvijay, A. K. D., & Dhanvijay, A. K. (2023). Large language models in hematology case solving: a comparative study of ChatGPT-3.5, Google Bard, and Microsoft Bing. Cureus, 15(8). doi: 10.7759/cureus.43861.

27. Dhanvijay, A. K. D., Pinjar, M. J., Dhokane, N., Sorte, S. R., Kumari, A., Mondal, H., & Dhanvijay, A. K. (2023). Performance of large language models (ChatGPT, Bing Search, and Google Bard) in solving case vignettes in physiology. Cureus, 15(8). doi: 10.7759/cureus.42972.

28. Al-Ashwal, F. Y., Zawiah, M., Gharaibeh, L., Abu-Farha, R., & Bitar, A. N. (2023). Evaluating the Sensitivity, Specificity, and Accuracy of ChatGPT-3.5, ChatGPT-4, Bing AI, and Bard Against Conventional Drug-Drug Interactions Clinical Tools. Drug, Healthcare and Patient Safety, 137–147. doi: 10.2147/DHPS.S425858.

29. Guillaume Cabanac, comment on PubPeer, https://pubpeer.com/publications/83DCF77815DC61C4ED6DCD88847EC4#1 (accessed on 11.10.2023)

30. Retraction Watch https://retractionwatch.com/2023/10/06/signs-of-undeclared-chatgpt-use-in-papers-mounting/ (accessed on 11.10.2023)

