## Supplementary File 1 for "Comparison of the audiological knowledge of three chatbots – ChatGPT, Bing Chat, and Bard"

Responses provided by ChatGPT. Errors are marked in red, inaccuracies in yellow.

<table>
<tr><th>No</th><th>Question</th><th>Chatbot response</th></tr>
<tr><td>1</td><td>What is an audiometric hearing test?</td><td>An audiometric hearing test, often simply referred to as an "audiogram," is a diagnostic procedure used to assess an individual's hearing ability. It is conducted by a trained audiologist or a licensed hearing healthcare professional and is a critical tool for diagnosing hearing loss and determining the degree and type of hearing impairment a person may have.<br>Here's how an audiometric hearing test typically works:<br>1. Preparation: Before the test begins, the audiologist will discuss your medical history and any specific concerns you may have about your hearing. They may also inquire about any exposure to loud noises or other factors that could contribute to hearing loss.<br>2. Physical Examination: The audiologist may conduct a visual inspection of your ears to check for any visible issues, such as earwax buildup or ear canal abnormalities, which can affect hearing.<br>3. Pure Tone Audiometry: This is the core component of the hearing test. You will be seated in a soundproof booth or room and given a pair of headphones or earphones. You'll be asked to listen for faint sounds at various frequencies (pitches) and signal when you hear them by pressing a button or raising your hand. This is usually done for both ears separately. The results are plotted on an audiogram, which is a graph that displays your hearing threshold levels at different frequencies.<br>o The horizontal axis of the audiogram represents frequency, typically measured in Hertz (Hz), ranging from low frequencies on the left to high frequencies on the right.<br>o The vertical axis represents the loudness or intensity of the sounds, usually measured in decibels (dB), with lower values at the top and higher values at the bottom.<br>4. Speech Audiometry: In some cases, the audiologist may also assess your ability to understand speech by playing recorded speech sounds or words at different volume levels. This helps determine the degree of speech discrimination or comprehension you have.<br>5. Bone Conduction Testing: If necessary, bone conduction testing may be performed. This involves placing a bone vibrator behind the ear to bypass the outer and middle ear and directly stimulate the inner ear. It helps differentiate between conductive and sensorineural hearing loss.<br>6. Interpretation: Based on the audiogram results, the audiologist can diagnose the type and degree of hearing loss. There are three main types of hearing loss:<br>o Conductive Hearing Loss: Caused by problems in the outer or middle ear, such as earwax blockage or middle ear infections.<br>o Sensorineural Hearing Loss: Caused by issues in the inner ear (cochlea) or the auditory nerve. This type is often permanent and may be age-related or due to exposure to loud noise.<br>o Mixed Hearing Loss: A combination of both conductive and sensorineural hearing loss.<br>7. Recommendations: After the test, the audiologist will discuss the results with you and may recommend further evaluations, hearing aids, medical treatment, or other interventions based on your specific hearing needs.<br>Audiometric hearing tests are essential for diagnosing hearing problems, understanding their nature, and determining the best course of action for managing them. Regular hearing tests are especially important as early detection and intervention can help prevent further hearing deterioration and improve the quality of life for individuals with hearing loss.</td></tr>
</table>

<table>
<tr><td>2</td><td>What is cochlear implant surgery?</td><td>A cochlear implant surgery is a medical procedure designed to treat severe to profound sensorineural hearing loss by bypassing damaged or non-functioning parts of the inner ear (cochlea) and directly stimulating the auditory nerve. Cochlear implants are electronic devices that can provide a sense of sound to individuals who do not benefit from traditional hearing aids.<br>Here's an overview of the cochlear implant surgery process:<ol><li>Candidacy Evaluation: Before considering cochlear implant surgery, a thorough evaluation is conducted by a team of medical professionals, including audiologists, otolaryngologists (ear, nose, and throat specialists), and speech-language pathologists. Candidates for cochlear implants typically have severe to profound sensorineural hearing loss, have limited benefit from hearing aids, and may have difficulty understanding speech even with amplification.</li><li>Implant Selection: If the candidate is deemed suitable for a cochlear implant, they and their healthcare team select an appropriate device and processor. Cochlear implants consist of an external sound processor worn behind the ear and an internal implant placed surgically within the ear.</li><li>Surgical Procedure:<ul><li>Anesthesia: The surgery is typically performed under general anesthesia.</li><li>Incision: An incision is made behind the ear to access the mastoid bone.</li><li>Internal Device Placement: A small opening is made in the mastoid bone, and the internal part of the cochlear implant is inserted into the cochlea. The electrodes are carefully threaded into the cochlea.</li><li>Connection: The internal device is then connected to a receiver-stimulator placed under the skin behind the ear. This receiver-stimulator picks up signals from the external sound processor and converts them into electrical impulses to stimulate the auditory nerve.</li><li>Wound Closure: The incision is closed, and the surgical site is dressed.</li></ul></li><li>Activation and Programming: After a healing period (usually a few weeks), the external sound processor is fitted and activated by an audiologist. The device is programmed specifically for the individual's hearing needs, with adjustments made over time to optimize their hearing experience.</li><li>Rehabilitation and Training: Cochlear implant recipients often undergo auditory training and rehabilitation to adapt to the new way of hearing. This may involve speech therapy and practice in understanding and interpreting the sounds provided by the implant.</li></ol>Cochlear implant surgery is considered safe and highly effective for suitable candidates, and it can significantly improve their ability to hear and understand speech. However, it's important to note that cochlear implants do not restore normal hearing but rather provide access to sound. The degree of improvement varies among individuals and depends on factors such as the duration of deafness, the age of implantation, and individual auditory processing abilities.<br>Cochlear implant surgery has transformed the lives of many people with severe hearing loss, allowing them to communicate more effectively and participate more fully in everyday life, including conversations, music enjoyment, and social interactions.</td></tr>
<tr><td>3</td><td>What is newborn hearing screening?</td><td>Newborn hearing screening, also known as neonatal hearing screening or infant hearing screening, is a medical procedure conducted shortly after birth to identify infants who may have hearing loss or hearing-related issues. The goal of newborn hearing screening is to detect hearing impairments early, ideally within the first few weeks of life, so that appropriate interventions can be initiated promptly to support the child's language and communication development.<br>There are two primary methods used for newborn hearing screening:</td></tr>
</table>

| | | |
|---|---|---|
| | | 1. **Otoacoustic Emissions (OAE) Testing**: In OAE testing, a small, soft earpiece or probe is placed in the baby's ear canal. Sounds or clicks are played through the earpiece, and the probe records the response from the baby's inner ear. If the baby's ear responds to the sounds appropriately, it indicates that the inner ear (cochlea) is functioning normally. If there is no response or a weak response, further evaluation may be necessary.<br>2. **Auditory Brainstem Response (ABR) Testing**: ABR testing involves placing electrodes on the baby's scalp, forehead, and behind the ears. Soft clicks or tones are played into the baby's ears through small earphones, and the electrodes measure the electrical activity of the auditory nerve and brainstem in response to the sounds. ABR testing can provide information about the functioning of the entire auditory pathway, from the ear to the brain.<br>The newborn hearing screening process is typically quick and painless, and it can be performed while the baby is asleep or resting. If a baby does not pass the initial hearing screening, it does not necessarily mean they have permanent hearing loss. There can be various reasons for a "refer" result, such as fluid in the ear canal, debris, or temporary hearing issues. Therefore, it is important for babies who do not pass the initial screening to undergo further evaluation.<br>When a baby does not pass the initial hearing screening, follow-up diagnostic testing is usually recommended. This more comprehensive evaluation helps determine the type and degree of hearing loss, if present, and provides valuable information for early intervention and management. Early intervention can include fitting the child with hearing aids, providing speech and language therapy, and offering support and resources to the family to help ensure the child's optimal development.<br>Newborn hearing screening programs are widely implemented in many countries as a standard practice to identify and address hearing issues early in a child's life. Timely detection and intervention can significantly improve a child's language and communication skills, social and academic development, and overall quality of life. |
| 4 | What is air-bone gap? | An air-bone gap is a term used in audiology to describe the difference in hearing thresholds between two types of hearing tests: air conduction and bone conduction. This gap in hearing thresholds indicates how well sound is transmitted through the air (using headphones or earphones) compared to how well it is transmitted through bone (typically through the skull bones). The air-bone gap is an essential concept in the evaluation of hearing loss and can provide valuable information about the location and nature of the hearing impairment.<br>Here's how air conduction and bone conduction tests work:<br>1. **Air Conduction**: This test involves the use of headphones or earphones to present sounds to the outer ear. The sounds travel through the ear canal, middle ear, and inner ear before reaching the cochlea, where they are processed and transmitted to the brain via the auditory nerve. The audiologist measures the softest sounds (thresholds) that the individual can hear at different frequencies (pitches) during the air conduction test.<br>2. **Bone Conduction**: In this test, a bone vibrator or oscillator is placed on the bone behind the ear (typically the mastoid bone). Vibrations generated by the bone vibrator bypass the outer and middle ear and directly stimulate the cochlea. The audiologist measures the softest bone-conducted sounds that the individual can hear at different frequencies.<br>The presence of an air-bone gap in a hearing test indicates that there is a difference in hearing sensitivity between the two methods. The specific characteristics of the gap can provide valuable diagnostic information:<br>• **Normal Hearing**: If there is no significant difference between air conduction and bone conduction thresholds, it suggests that the individual has normal hearing.<br>• **Conductive Hearing Loss**: When air conduction thresholds are significantly worse than bone conduction thresholds, it indicates a conductive hearing loss. This means that sound is not being conducted effectively through the outer and |

| | | |
|---|---|---|
| | | middle ear to reach the inner ear. Common causes of conductive hearing loss include earwax blockage, middle ear infections, or problems with the ear ossicles (small bones in the middle ear).<br>• **Sensorineural Hearing Loss**: If both air conduction and bone conduction thresholds are elevated, but the gap remains consistent, it suggests a sensorineural hearing loss. This type of hearing loss originates in the inner ear or along the auditory nerve. Common causes include age-related hearing loss, exposure to loud noise, and genetic factors.<br>• **Mixed Hearing Loss**: A mixed hearing loss occurs when there is both a sensorineural component (elevated air conduction thresholds) and a conductive component (elevated bone conduction thresholds), resulting in an air-bone gap. This suggests that there is a problem in both the inner and middle ear.<br>In summary, the air-bone gap is a critical concept in audiology used to differentiate between conductive and sensorineural hearing loss and to guide the diagnosis and treatment of hearing disorders. It helps audiologists determine the most appropriate interventions, such as hearing aids, surgical procedures, or medical management, based on the underlying causes of the hearing impairment. |
| 5 | What is auditory neuropathy and how is it diagnosed? | Auditory neuropathy, also known as auditory neuropathy spectrum disorder (ANSD), is a hearing disorder that affects the transmission of sound signals from the inner ear (cochlea) to the brain through the auditory nerve. Unlike typical sensorineural hearing loss, where the cochlea is responsible for the hearing impairment, in auditory neuropathy, the problem lies in the transmission of signals along the auditory nerve or the synapses connecting the cochlea and the auditory nerve. This can result in difficulties in processing and understanding speech, even though sounds may be detected.<br>Diagnosing auditory neuropathy involves a combination of hearing tests and assessments to evaluate the specific nature of the auditory disorder. Here are the steps involved in diagnosing auditory neuropathy:<br>1. **Pure Tone Audiometry**: The initial evaluation typically includes pure tone audiometry (air conduction and bone conduction tests). This helps determine the degree and type of hearing loss. Individuals with auditory neuropathy may show a wide range of hearing thresholds, from normal to profound, on pure tone audiometry.<br>2. **Tympanometry**: Tympanometry assesses the function of the middle ear, specifically the mobility of the eardrum and the condition of the middle ear. In some cases, individuals with auditory neuropathy may have normal middle ear function, but this test helps rule out conductive hearing loss.<br>3. **Otoacoustic Emissions (OAE)**: OAE testing assesses the response of the outer hair cells in the cochlea to sound. People with auditory neuropathy may have normal OAE responses, indicating that their cochlea is functioning normally in terms of amplifying sounds.<br>4. **Auditory Brainstem Response (ABR)**: ABR testing measures the electrical activity of the auditory nerve and the brainstem in response to sound stimuli. In individuals with auditory neuropathy, ABR results typically show abnormal or absent responses, suggesting a problem with the transmission of signals from the cochlea to the brainstem.<br>5. **Speech Audiometry**: Speech audiometry involves testing the individual's ability to understand speech sounds at different volumes. Individuals with auditory neuropathy may have difficulty understanding speech, even when their pure tone audiometry results indicate some degree of hearing.<br>6. **Auditory Steady-State Response (ASSR) Testing**: This is an advanced auditory assessment that may be used to further evaluate the individual's hearing and auditory nerve function. ASSR can provide more detailed information about the specific frequencies and hearing thresholds affected by auditory neuropathy. |

| | | |
|---|---|---|
| | | 7. **Medical Evaluation**: In some cases, a medical evaluation may be performed to rule out underlying medical conditions that could contribute to auditory neuropathy.<br>Diagnosing auditory neuropathy can be complex, and a combination of these tests is often necessary to confirm the condition. Once diagnosed, the management of auditory neuropathy may include interventions such as hearing aids, cochlear implants, or auditory training. The appropriate treatment plan depends on the individual's specific hearing needs and the degree of impairment.<br>It's essential to consult with an audiologist or an ear, nose, and throat specialist (otolaryngologist) if you suspect auditory neuropathy or have concerns about hearing difficulties, especially if traditional hearing tests do not provide a clear diagnosis. Early intervention and appropriate management can significantly improve a person's ability to communicate and interact effectively in their environment. |
| 6 | Does the stapedius reflex protect the inner ear from damage? | Yes, the stapedius reflex, also known as the acoustic reflex, plays a role in protecting the inner ear from loud and potentially damaging sounds. It is one of the body's natural defense mechanisms against excessive noise exposure.<br>The stapedius reflex involves the contraction of the stapedius muscle, which is one of the muscles of the middle ear. The stapedius muscle is attached to the stapes bone, one of the tiny bones (ossicles) in the middle ear. When exposed to a loud sound, the auditory system detects the sound's intensity, and if it reaches a certain threshold, the stapedius muscle contracts involuntarily.<br>Here's how the stapedius reflex helps protect the inner ear from damage:<br>1. **Sound Dampening**: When the stapedius muscle contracts, it pulls the stapes bone away from the oval window of the cochlea (the fluid-filled part of the inner ear responsible for hearing). This action reduces the transmission of sound vibrations from the middle ear to the inner ear, effectively dampening the intensity of the sound reaching the delicate hair cells within the cochlea.<br>2. **Protection from Loud Noises**: The stapedius reflex is particularly sensitive to loud and sudden sounds. By dampening the intensity of these sounds, it helps prevent overstimulation and potential damage to the hair cells within the cochlea. Hair cells are essential for converting sound vibrations into electrical signals that the brain can interpret as sound.<br>While the stapedius reflex provides some level of protection against loud noises, it has limitations. It is not an instantaneous response, and there may still be some exposure to loud sounds before the reflex fully engages. Additionally, the reflex may not be as effective against continuous or prolonged loud noises as it is against sudden and brief sounds.<br>In situations of chronic or extreme noise exposure, the stapedius reflex may become less effective, and repeated exposure to loud sounds can still lead to noise-induced hearing loss. Therefore, it is essential to use additional protective measures, such as wearing hearing protection (e.g., earplugs or earmuffs) in noisy environments, to further safeguard the inner ear from damage caused by excessive noise.<br>Overall, the stapedius reflex is a valuable natural defense mechanism that contributes to the protection of the inner ear from loud sounds, but it should not be relied upon as the sole means of hearing protection in noisy environments. |
| 7 | What are the clinical signs of conductive hearing loss? | Conductive hearing loss is a type of hearing impairment that occurs when sound waves are unable to travel effectively through the outer or middle ear to reach the inner ear (cochlea). This type of hearing loss can result from various underlying causes or conditions that disrupt the normal transmission of sound. Clinical signs of conductive hearing loss can vary depending on its severity and the specific underlying cause, but some common signs and symptoms include: |

| | | |
|---|---|---|
| | | 1. **Reduced Hearing**: The primary and most noticeable sign of conductive hearing loss is a decrease in hearing sensitivity. Individuals with this type of hearing loss often report that sounds seem muffled, quieter, or less clear.<br>2. **Difficulty Hearing Soft Sounds**: People with conductive hearing loss may have difficulty hearing faint or soft sounds, such as whispers, distant voices, or the rustling of leaves.<br>3. **Ear Fullness or Pressure**: Some individuals with conductive hearing loss experience a sensation of fullness or pressure in the affected ear. This sensation may be due to underlying conditions like earwax impaction, ear infections, or eustachian tube dysfunction.<br>4. **Tinnitus**: Tinnitus refers to the perception of ringing, buzzing, or other noises in the ear when no external sound source is present. Conductive hearing loss can sometimes be associated with tinnitus, and individuals may report hearing these sounds more prominently.<br>5. **Ear Pain or Discomfort**: Depending on the underlying cause of conductive hearing loss, individuals may experience ear pain or discomfort. This can occur with conditions like ear infections or when there is a blockage in the ear canal.<br>6. **Sensation of "Plugged" Ears**: People with conductive hearing loss may describe their ears as feeling "plugged" or as if something is blocking their ear canals.<br>7. **Difficulty Understanding Speech**: While conductive hearing loss primarily affects the volume of sounds, it can also impact the clarity of speech. Speech may sound unclear or distorted to individuals with conductive hearing loss.<br>8. **Tendency to Speak Loudly**: Individuals with conductive hearing loss may unintentionally speak louder than normal because they perceive their own voice as being quieter than it actually is.<br>9. **Fluid Drainage**: In cases of conductive hearing loss due to middle ear infections (otitis media), there may be visible fluid drainage from the ear.<br>10. **History of Ear Problems**: A significant number of people with conductive hearing loss have a history of recurrent ear infections, ear surgeries, or other ear-related issues.<br>It's important to note that the specific signs and symptoms of conductive hearing loss can vary depending on the underlying cause. Some cases of conductive hearing loss may be temporary and easily treatable, while others may require medical intervention, such as surgery or the use of hearing aids. If you or someone you know is experiencing any of these signs or symptoms, it's essential to seek evaluation and care from a qualified healthcare provider or audiologist to determine the cause and appropriate treatment. |
| 8 | Does it make sense to do a auditory brainstem responses examination if an MRI confirmed large cerebello pontine angle tumor in one ear? | If an MRI has already confirmed the presence of a large cerebellopontine angle (CPA) tumor in one ear, it may not be necessary to perform an auditory brainstem response (ABR) examination to evaluate hearing in that specific ear. The MRI findings, along with clinical information and a patient's medical history, provide a clear indication of the tumor's location and its potential impact on auditory structures in the CPA.<br>An ABR test is typically conducted to assess the function of the auditory nerve and brainstem, primarily when there is uncertainty about the cause of hearing loss or to investigate hearing issues in cases such as newborns or young children who cannot provide reliable feedback during a conventional hearing assessment.<br>In the context of a known large CPA tumor, an ABR may not be necessary for several reasons:<br>1. **MRI Confirmation**: The MRI is a highly sensitive imaging technique that can precisely visualize the presence, size, and location of the tumor within the cerebellopontine angle. It can provide essential information about the tumor's relationship to nearby structures, including the cochlear and auditory nerve. |

| | | |
|---|---|---|
| | | 2. **Hearing Impact**: In cases where a large CPA tumor is detected and is in proximity to the auditory nerve, it is highly likely that the tumor is responsible for any hearing loss in the affected ear. The tumor's compression or damage to the auditory nerve or associated structures can explain the hearing impairment.<br>3. **Treatment Planning**: Once the tumor has been identified through MRI, the focus typically shifts to treatment planning, which may involve surgical intervention (e.g., tumor resection), radiation therapy, or other medical management. The goal is often to address the tumor's underlying cause and, if possible, preserve or restore hearing.<br>4. **Risks and Benefits**: Performing an ABR test in the presence of a large CPA tumor may carry certain risks, such as discomfort or potential complications related to the testing procedure. It is essential to weigh these potential risks against the expected benefits, especially when the primary goal is to address the tumor itself.<br>In cases of hearing loss associated with large CPA tumors, the emphasis should be on the comprehensive management of the tumor and the preservation or restoration of hearing, rather than solely on confirming the hearing impairment through additional testing. The treating healthcare team, which may include neurosurgeons, otologists, and audiologists, will work together to determine the most appropriate diagnostic and treatment plan for the individual's specific situation.<br>It's crucial for individuals with CPA tumors to have a thorough evaluation and ongoing medical care, as these tumors can have various effects on hearing and other neurological functions. Consultation with specialists and a comprehensive treatment plan should guide the management of these complex cases. |
| 9 | What is the therapeutic management of acoustic neuroma? | The therapeutic management of an acoustic neuroma, also known as a vestibular schwannoma, involves a combination of treatment approaches aimed at controlling the growth of the tumor, preserving hearing and other neurological functions, and addressing symptoms. The choice of treatment depends on various factors, including the size of the tumor, the patient's age and overall health, and the individual's hearing status. The main treatment options for acoustic neuromas include:<br>1. **Observation (Watchful Waiting):** For small tumors that are not causing significant symptoms and do not pose an immediate threat to hearing or nearby structures, a strategy of observation may be recommended. Regular MRI scans are performed to monitor tumor growth over time. This approach is often chosen for elderly patients or those with significant medical comorbidities who may not be suitable candidates for more aggressive treatments.<br>2. **Surgery:**<br>o **Microsurgical Resection (Translabyrinthine or Retrosigmoid Approach):** Surgery involves the removal of the tumor through one of these approaches, depending on factors such as tumor size and location. Microsurgery aims to preserve facial nerve function, hearing (if possible), and other neurological functions while completely removing the tumor. The choice of surgical approach and the extent of tumor removal depend on individual circumstances.<br>o **Facial Nerve Monitoring:** During surgery, facial nerve function is closely monitored to minimize the risk of damage to the facial nerve, which controls facial movements. Nerve monitoring helps the surgeon make real-time adjustments to protect the facial nerve.<br>o **Hearing Preservation Surgery:** In select cases, particularly for small tumors and patients with good hearing, surgical techniques may be used to attempt to preserve or improve hearing. This is known as hearing preservation surgery.<br>3. **Radiation Therapy:**<br>o **Stereotactic Radiosurgery (e.g., Gamma Knife, CyberKnife):** This non-invasive procedure delivers focused beams of radiation to the tumor, typically over multiple sessions. The goal is to stop tumor growth |

| | | |
|---|---|---|
| | | or shrink the tumor over time. Radiosurgery is often considered for small to medium-sized tumors or for patients who are not surgical candidates.<br>o **Fractionated Stereotactic Radiotherapy:** In some cases, radiation therapy is delivered in smaller, fractionated doses over several weeks. This approach may be used for larger tumors or when preserving hearing is a priority.<br>4. **Combination Therapy:** In certain situations, a combination of treatments may be recommended, such as surgery followed by radiation therapy (either immediately or as a secondary treatment) to address any residual tumor or prevent regrowth.<br>5. **Supportive and Symptom Management:** Regardless of the chosen treatment approach, patients may require supportive care and symptom management. This can include physical therapy for balance issues, hearing rehabilitation, and psychological support to address the emotional and psychological impact of the diagnosis and treatment.<br>The specific treatment plan for each patient is determined through careful evaluation by a team of healthcare professionals, which may include neurosurgeons, otolaryngologists (ear, nose, and throat specialists), radiation oncologists, and audiologists. Treatment decisions take into account the patient's overall health, tumor characteristics, hearing status, and individual preferences.<br>Regular follow-up with MRI scans is typically necessary to monitor for tumor recurrence or regrowth, especially for patients who undergo conservative management or radiation therapy. The goal of therapeutic management for acoustic neuromas is to achieve the best possible outcome while preserving hearing and minimizing complications. |
| 10 | What is hidden hearing loss? | Hidden hearing loss, also known as "hidden hearing deficit" or "cochlear synaptopathy," is a relatively recent and somewhat complex concept in the field of audiology and hearing science. Unlike traditional forms of hearing loss, which are typically associated with reduced sensitivity to sounds (measured using pure-tone audiometry), hidden hearing loss is characterized by difficulties in processing and understanding speech, particularly in noisy or challenging listening environments, even when standard audiometric tests indicate normal or near-normal hearing thresholds.<br>Key characteristics and aspects of hidden hearing loss include:<br>1. **Normal Pure-Tone Audiogram:** Individuals with hidden hearing loss typically have normal or near-normal hearing thresholds when tested using pure-tone audiometry. This means they can hear soft sounds at various frequencies (pitches) as well as or even better than individuals with typical hearing.<br>2. **Difficulty Understanding Speech:** Despite having normal hearing thresholds, individuals with hidden hearing loss often report difficulty understanding speech, especially in situations with background noise or in environments with multiple competing sound sources. This difficulty in speech perception can lead to communication challenges, particularly in social and noisy settings.<br>3. **Synaptopathy and Neural Damage:** The underlying mechanism of hidden hearing loss is believed to involve damage or dysfunction in the synapses (connections) between inner hair cells and auditory nerve fibers in the cochlea. This damage may result from various factors, including noise exposure, aging, and certain medical conditions.<br>4. **Auditory Processing Deficits:** Research suggests that individuals with hidden hearing loss may exhibit deficits in central auditory processing, affecting their ability to discriminate and process complex auditory information. This can contribute to speech perception difficulties in noisy environments. |

| | | |
|---|---|---|
| | | 5. **Subclinical Hearing Loss:** While standard audiometric tests focus on the detection of hearing loss based on sensitivity to pure tones, hidden hearing loss may be described as a form of "subclinical" hearing loss, meaning it is not readily detectable using these tests.<br>It's important to note that the term "hidden hearing loss" is still a topic of ongoing research, and there is some debate and variability in its definition and diagnosis within the scientific community. Researchers are working to better understand the underlying mechanisms and refine diagnostic criteria for this condition.<br>Treatment and management options for hidden hearing loss are also an area of active investigation. Some potential interventions may include auditory training and rehabilitation programs to improve speech perception in challenging listening environments.<br>If you suspect you have hidden hearing loss or are experiencing difficulties understanding speech despite normal hearing thresholds on standard hearing tests, it is advisable to consult with an audiologist or hearing healthcare professional. They can conduct more specialized assessments and provide guidance on potential management strategies tailored to your specific needs. |
