## Supplementary File 2 for "Comparison of the audiological knowledge of three chatbots – ChatGPT, Bing Chat, and Bard"

Responses provided by Bing Chat. Errors are marked in red, inaccuracies in yellow. 'Dowiedz się więcej:' – means learn more, and comes from Polish version of Bing Chat.

| No | Question | Bing Chat response |
|---|---|---|
| 1 | What is an audiometric hearing test? | An audiometric hearing test is a noninvasive hearing test that measures a person's ability to hear different sounds and pitches. It tests both the intensity and the tone of sounds, balance issues, and other issues related to the function of the inner ear. A doctor who specializes in diagnosing and treating hearing loss called an audiologist administers the test. The unit of measure for sound intensity is the decibel (dB). A healthy human ear can hear quiet sounds such as whispers. These are about 20 dB. A loud sound such as a jet engine is between 140 and 180 dB. The tone of a sound is measured in cycles per second. The unit of measure for tone is Hertz (Hz). Humans can hear tones between 20-20,000 Hz. An audiometry exam requires no special preparation. All you have to do is show up to your appointment on time and follow the audiologist's instructions[1].<br>Dowiedz się więcej:<br>1. https://www.healthline.com/health/audiology<br>2. https://www.hearingsol.com/help/hearing-test/types/audiometry/<br>3. https://www.healthline.com/health/audiogram<br>4. https://en.wikipedia.org/wiki/Audiometry |
| 2 | What is cochlear implant surgery? | Cochlear implant surgery is a fairly routine surgery that is typically performed under general anesthesia. The surgeon will make a small incision behind the ear and sometimes a small area of hair may be shaved away from the incision site. The implant is then placed under the skin and the electrode is inserted into the inner ear. Cochlear implants can improve hearing in people with severe hearing loss who are no longer helped by using hearing aids. Cochlear implants can improve their communication and quality of life. Cochlear implants may be placed in one ear (unilateral) or both ears (bilateral)[12].<br>Dowiedz się więcej:<br>1. https://www.mayoclinic.org/tests-procedures/cochlear-implants/about/pac-20385021<br>2. https://en.wikipedia.org/wiki/Cochlear_implant<br>3. https://www.cochlear.com/us/en/home/diagnosis-and-treatment/implant-preparation-and-procedures/cochlear-implant-surgery |
| 3 | What is newborn hearing screening? | Newborn hearing screening is a test that helps identify babies who have permanent hearing loss as early as possible. This means parents can get the support and advice they need right from the start. Hearing loss in babies 1 to 2 babies in every 1,000 are born with permanent hearing loss in 1 or both ears. This increases to about 1 in every 100 babies who have spent more than 48 hours in intensive care. Most of these babies are born into families with no history of permanent hearing loss. Permanent hearing loss can significantly affect babies' development. Finding out early can give these babies a better chance of developing language, speech and communication skills. It will also help them make the most of relationships with their family or carers from an early age. The newborn hearing test is called the automated otoacoustic emission (AOAE) test. It takes just a few minutes. A small soft-tipped earpiece is placed in your baby's ear and gentle clicking sounds are played[1].<br>Dowiedz się więcej:<br>1. https://www.nhs.uk/conditions/baby/newborn-screening/hearing-test/<br>2. https://iris.who.int/bitstream/handle/10665/339288/9789241599496-eng.pdf<br>3. https://www.cdc.gov/ncbddd/hearingloss/parentsguide/understanding/newbornhearingscreening.html |
| 4 | What is air-bone gap? | Air-bone gap is the difference between the air conduction threshold and the bone conduction threshold of a patient's hearing. It is used to determine the type of hearing loss that a patient has. If the air-bone gap is greater than 10 decibels (dB), then the patient has conductive hearing loss. If the air-bone gap is less than or equal to 10 dB, then the patient has sensorineural hearing loss. |

| | | |
|---|---|---|
| 5 | What is auditory neuropathy and how is it diagnosed? | Auditory neuropathy is a hearing disorder in which the inner ear successfully detects sound, but has a problem with sending sound from the ear to the brain. It can affect people of all ages, from infancy through adulthood. The number of people affected by auditory neuropathy is not known, but current information suggests that auditory neuropathies play a substantial role in hearing impairments and deafness. When their hearing sensitivity is tested, people with auditory neuropathy may have normal hearing or hearing loss ranging from mild to severe. They always have poor speech-perception abilities, meaning that they have trouble understanding speech clearly. People with auditory neuropathy have greater impairment in speech perception than hearing health experts would predict based upon their degree of hearing loss on a hearing test[1]. Health professionals—including otolaryngologists (ear, nose, and throat doctors), pediatricians, and audiologists—use a combination of methods to diagnose auditory neuropathy. These include tests of auditory brainstem response (ABR) and otoacoustic emissions (OAE)[1][2].<br>Dowiedz się więcej:<br>1. https://www.nidcd.nih.gov/health/auditory-neuropathy<br>2. https://en.wikipedia.org/wiki/Auditory_neuropathy<br>3. https://audiologyisland.com/blog/hearing-issues-treatment-what-should-you-know-about-auditory-neuropathy-spectrum-disorder/<br>4. https://www.betterhealth.vic.gov.au/health/conditionsandtreatments/hearing-loss-auditory-neuropathy |
| 6 | Does the stapedius reflex protect the inner ear from damage? | Yes, the stapedius muscle is involved in the auditory middle ear reflex and has a crucial role in protecting the auditory system from damage1. The stapedius muscle is innervated by the stapedial branch of facial nerve[1]. When the incoming sound is loud enough to stimulate the receptor cells in the inner ear, the afferent signal reaches the cochlear nucleus in the brainstem via vestibulocochlear nerve (CN VIII). From the brainstem, efferent signals are sent to the ipsilateral and contralateral middle ears triggering the contractions of the stapedius muscles. Their contractions result with posterior rotation and placing the base of the stapes into the oval window which closes it and attenuates further vibrations passed to cochlea. Thus, the main function of the stapedius muscle is to protect the hearing apparatus when exposed to loud sounds[1].<br>I hope this helps!<br>Dowiedz się więcej:<br>1. https://www.kenhub.com/en/library/anatomy/stapedius-muscle<br>2. https://www.verywellhealth.com/stapedius-5498666<br>3. https://www.hearingresearch.org/acoustic-reflex/ |
| 7 | What are the clinical signs of conductive hearing loss? | Some of the common signs of conductive hearing loss include sudden or unexpected hearing loss in one or both ears, all sounds seeming muffled or blocked in one ear, full or “stuffy” sensation in the ear, dizziness, draining of the ear, and pain or tenderness in the ear[1][2]. However, symptoms can vary depending on the exact cause and severity[1].<br>I hope this helps!<br>Dowiedz się więcej:<br>1. https://www.enthealth.org/conditions/conductive-hearing-loss/<br>2. https://www.verywellhealth.com/conductive-hearing-loss-5225503<br>3. https://myhearingcenters.com/blog/conductive-hearing-loss-signs-causes-and-treatments/<br>4. https://www.forbes.com/health/hearing-aids/conductive-hearing-loss/<br>5. https://www.houstonent.com/blog/what-is-conductive-hearing-loss-causes-symptoms-and-treatment-options |
| 8 | Does it make sense to do a auditory brainstem responses | An auditory brainstem response (ABR) examination is a test that measures the electrical activity of the auditory nerve and the brainstem in response to sound[1]. It is used to evaluate hearing loss and other disorders of the auditory nerve and brainstem. |

| | | |
|---|---|---|
| | examination if an MRI confirmed large cerebello pontine angle tumor in one ear? | However, if an MRI has confirmed a large cerebellopontine angle tumor in one ear, it is best to consult with your doctor or audiologist to determine if an ABR examination is appropriate for you[2].<br>I hope this helps!<br>Dowiedz się więcej:<br>1. https://www.cambridge.org/core/journals/journal-of-laryngology-and-otology/article/abs/brain-stem-electrical-responses-in-cerebellopontine-angle-tumours/F9A0DD136AA56543727AC007908C6793<br>2. https://radiopaedia.org/articles/cerebellopontine-angle-mass<br>3. https://www.cambridge.org/core/journals/journal-of-laryngology-and-otology/article/abs/audiovestibular-findings-in-meningioma-of-the-cerebellopontine-angle-a-retrospective-review/B5780D2E1E3EEDA36F8D35A2E24CAC28 |
| 9 | What is the therapeutic management of acoustic neuroma? | The therapeutic management of acoustic neuroma depends on the size and location of the tumor, as well as the patient's age and overall health. Treatment options include observation, surgery, and radiation therapy. The goal of treatment is to remove or control the tumor while preserving hearing and other neurological functions.<br>I hope this helps! |
| 10 | What is hidden Hearing loss? | Hidden hearing loss is a type of hearing loss that can't be detected by some hearing tests. It is a result of disconnection in sound transmission between your inner ear and brain[1]. Unlike other types of hearing loss, hidden hearing loss happens when there is a defect in your cochlea, a bone in the inner ear[1]. Factors such as old age, exposure to loud noise, and taking certain drugs can increase your risk of developing hidden hearing loss[1].<br>I hope this helps!<br>Dowiedz się więcej:<br>1. https://www.verywellhealth.com/hidden-hearing-loss-5218694<br>2. https://www.healthyhearing.com/report/53066-Hidden-hearing-loss |
