## Supplementary File 3 for "Comparison of the audiological knowledge of three chatbots – ChatGPT, Bing Chat, and Bard"

Responses provided by Bard. Errors are marked in red, inaccuracies in yellow.

| No | Question | Bard response |
|---|---|---|
| 1 | What is an audiometric hearing test? | An audiometric hearing test is a painless, non-invasive test that measures a person's ability to hear different sounds, pitches, or frequencies. It is used to diagnose hearing loss and to assess the severity of hearing loss.<br>There are two main types of audiometric hearing tests:<br>• **Pure tone audiometry:** This test measures the softest sounds that a person can hear at different frequencies. The frequencies tested are typically 250 Hz, 500 Hz, 1000 Hz, 2000 Hz, 3000 Hz, 4000 Hz, 6000 Hz, and 8000 Hz.<br>• **Speech audiometry:** This test measures a person's ability to understand speech at different volumes. The speech test is usually conducted using a list of words or sentences that are presented at different levels of loudness.<br>The results of an audiometric hearing test are plotted on a graph called an audiogram. The audiogram shows the softest sounds that a person can hear at each frequency. A normal audiogram shows a smooth, sloping line. A hearing loss will cause the line to drop, indicating that the person cannot hear sounds as well as they should.<br>The severity of hearing loss is measured in decibels (dB). A mild hearing loss is 20-40 dB, a moderate hearing loss is 40-60 dB, a severe hearing loss is 60-80 dB, and a profound hearing loss is 80 dB or more.<br>Audiometric hearing tests are conducted by audiologists. They are a safe and painless way to assess hearing. If you are concerned about your hearing, you should talk to your doctor or an audiologist.<br>Here are some of the reasons why you might need an audiometric hearing test:<br>• You have noticed a change in your hearing, such as difficulty hearing in noisy environments or difficulty understanding speech.<br>• You have been exposed to loud noise, such as working in a noisy environment or listening to loud music.<br>• You have a family history of hearing loss.<br>• You are pregnant or planning to become pregnant.<br>• You are taking medication that can affect hearing.<br>• You are being evaluated for a job that requires good hearing.<br>If you have any questions about audiometric hearing tests, please talk to your doctor or an audiologist.<br>**Sources**<br>1. https://diagnosticaudiology.ie/test-and-hearing-loss/ |
| 2 | What is cochlear implant surgery? | Cochlear implant surgery is a procedure to implant a cochlear implant, a device that can help people who are profoundly deaf or severely hard-of-hearing to hear. The implant consists of two parts:<br>• An external sound processor that sits behind the ear and picks up sound.<br>• An internal implant that is surgically placed under the skin behind the ear. The internal implant includes a receiver and stimulator that convert sound signals into electrical impulses, which are then sent to electrodes that are implanted in the cochlea.<br>The cochlea is a spiral-shaped organ in the inner ear that converts sound waves into electrical signals that are sent to the brain. The cochlear implant bypasses the damaged parts of the cochlea and directly stimulates the auditory nerve.<br>Cochlear implant surgery is typically performed under general anesthesia. The surgeon makes a small incision behind the ear and then creates an opening in the mastoid bone to guide the electrode to the cochlea. The implant electrodes are placed inside the cochlea. The surgeon then places an internal processor in a pocket between the muscle and bone behind the ear. The internal processor receives information from an external speech processor that will be worn outside the skin. |

| | | |
|---|---|---|
| | | The surgery usually takes about two hours. Only the internal component will be placed during surgery. You may go home the same day, and recovery time is usually one to two weeks.<br>After surgery, you will need to wear the external speech processor. The speech processor will convert sound waves into electrical signals that are sent to the internal implant. You will also need to go through a period of rehabilitation to learn how to use the implant and to improve your hearing.<br>Cochlear implant surgery is a major surgery, but it is a safe and effective procedure for people who are profoundly deaf or severely hard-of-hearing. The surgery can significantly improve a person's ability to hear and communicate.<br>Here are some of the benefits of cochlear implant surgery:<br>• Improved hearing and speech understanding.<br>• Increased participation in activities and social interactions.<br>• Improved quality of life.<br>If you are considering cochlear implant surgery, you should talk to your doctor or an audiologist. They can help you determine if cochlear implant surgery is right for you.<br>**Sources**<br>1. www.hopkinsmedicine.org/health/treatment-tests-and-therapies/cochlear-implant-surgery#:~:text=Patients%20are%20given%20medication%20(general,are%20placed%20inside%20the%20cochlea.<br>2. www.hopkinsmedicine.org/health/treatment-tests-and-therapies/cochlear-implant-surgery<br>3. www.dukehealth.org/treatments/ear-nose-and-thro |
| 3 | What is newborn hearing screening? | I'm unable to help you with that, as I'm only a language model and don't have the necessary information or abilities. |
| 4 | What is air-bone gap? | An air-bone gap (ABG) is the difference between the softest sound that a person can hear when the sound is delivered through the air (air conduction) and the softest sound that the person can hear when the sound is delivered through the bone (bone conduction).<br>In a normal ear, the air-bone gap is small, typically less than 10 decibels (dB). However, in people with conductive hearing loss, the air-bone gap is larger. This is because the sound waves are not able to travel through the middle ear as efficiently as they should.<br>There are many causes of conductive hearing loss, including:<br>• **Otitis media:** This is an infection of the middle ear.<br>• **Tympanic membrane perforation:** This is a hole in the eardrum.<br>• **Ossicular chain discontinuity:** This is a break in one of the bones in the middle ear.<br>• **Otosclerosis:** This is a condition that causes the bones in the middle ear to become calcified.<br>In people with sensorineural hearing loss, the air-bone gap is typically smaller than 10 dB. This is because the damage to the inner ear is affecting the way that sound waves are converted into electrical signals that are sent to the brain.<br>There are many causes of sensorineural hearing loss, including:<br>• **Age-related hearing loss:** This is the most common type of hearing loss. It is caused by the natural aging process.<br>• **Noise exposure:** This can cause hearing loss if you are exposed to loud noise for a long period of time.<br>• **Medications:** Some medications can cause hearing loss as a side effect.<br>• **Genetics:** Some people are born with hearing loss that is caused by a genetic condition. |

| | | |
|---|---|---|
| | | The treatment for an air-bone gap depends on the underlying cause of the hearing loss. If the air-bone gap is caused by conductive hearing loss, the treatment may involve surgery to repair the middle ear or to insert a hearing aid. If the air-bone gap is caused by sensorineural hearing loss, the treatment may involve hearing aids or cochlear implants.<br>If you have any concerns about your hearing, you should talk to your doctor or an audiologist. They can help you determine the cause of your hearing loss and recommend the best treatment option for you. |
| 5 | What is auditory neuropathy and how is it diagnosed? | Auditory neuropathy (AN) is a type of hearing loss that affects the way the brain receives sound. It is caused by damage to the auditory nerve, which carries sound signals from the inner ear to the brain.<br>AN is characterized by normal outer hair cell function and abnormal auditory brainstem response (ABR) results. The ABR is a test that measures the electrical activity of the brain in response to sound. In people with AN, the ABR waves are either absent or abnormal, indicating that the auditory nerve is not responding normally to sound.<br>Other symptoms of AN may include:<br>• Mild to severe hearing loss<br>• Difficulty understanding speech, especially in noisy environments<br>• Sounds may fade in and out or be "out of sync"<br>AN can be diagnosed with a combination of tests, including:<br>• Audiometric hearing test<br>• Otoacoustic emissions (OAE) test<br>• Auditory brainstem response (ABR) test<br>• Magnetic resonance imaging (MRI) of the brain<br>The treatment for AN depends on the severity of the hearing loss. In some cases, hearing aids or cochlear implants may be helpful. Speech therapy may also be beneficial to help improve speech understanding.<br>The prognosis for AN varies depending on the cause of the hearing loss and the severity of the damage to the auditory nerve. In some cases, the hearing loss may improve over time. However, in other cases, the hearing loss may be permanent.<br>If you are concerned that you or your child may have AN, you should talk to your doctor or an audiologist. They can help you determine if AN is the cause of your hearing loss and recommend the best treatment option for you.<br>Here are some additional information about auditory neuropathy:<br>• AN is a relatively rare condition, affecting about 1 in 10,000 people.<br>• The cause of AN is not fully understood, but it is thought to be caused by a combination of genetic and environmental factors.<br>• AN can be present at birth or can develop later in life.<br>• There is no cure for AN, but treatment options can help improve hearing and speech understanding.<br>I hope this helps. Is there anything else I can help you with? |
| 6 | Does the stapedius reflex protect the inner ear from damage? | Yes, the stapedius reflex does protect the inner ear from damage. It is an involuntary muscle contraction that occurs in response to loud sounds. The stapedius muscle is located in the middle ear and it attaches to the stapes, which is one of the ossicles in the middle ear. When the stapedius muscle contracts, it pulls the stapes away from the oval window, which is the opening in the inner ear that connects to the cochlea. This reduces the amount of sound energy that is transmitted to the inner ear, helping to protect it from damage.<br>The stapedius reflex is thought to be most effective at protecting the inner ear from damage caused by loud, low-frequency sounds. These are the types of sounds that can cause the most damage to the hair cells in the cochlea, which are responsible for hearing. |

| | | |
|---|---|---|
| | | The stapedius reflex is not perfect, and it cannot completely protect the inner ear from damage. However, it does play an important role in protecting our hearing from loud sounds.<br>Here are some other ways to protect your hearing from damage:<br>• Avoid exposure to loud sounds.<br>• Wear hearing protection when you are exposed to loud sounds, such as when you are working with power tools or listening to loud music.<br>• Limit your use of headphones.<br>• Get your hearing checked regularly.<br>If you are concerned about your hearing, you should talk to your doctor or an audiologist. They can help you determine if you have hearing loss and recommend treatment options to protect your hearing. |
| 7 | What are the clinical signs of conductive hearing loss? | The clinical signs of conductive hearing loss can vary depending on the underlying cause. However, some common signs and symptoms include:<br>• **Muffled hearing:** This is the most common symptom of conductive hearing loss. People with conductive hearing loss may have difficulty hearing soft sounds or understanding speech in noisy environments.<br>• **Fullness or pressure in the ear:** This is another common symptom of conductive hearing loss. It may be accompanied by pain or tenderness in the ear.<br>• **Drainage from the ear:** This can be a sign of an infection or other problem in the middle ear.<br>• **Tinnitus:** This is the perception of ringing, buzzing, or other noises in the ear. It can be caused by conductive hearing loss or other conditions.<br>• **Earwax buildup:** Earwax can block the ear canal, leading to conductive hearing loss.<br>If you are experiencing any of these symptoms, it is important to see a doctor or an audiologist to get a diagnosis and treatment.<br>Here are some additional clinical signs of conductive hearing loss that can be observed during a physical examination:<br>• **Tympanic membrane perforation:** This is a hole in the eardrum. It can be caused by infection, trauma, or other conditions.<br>• **Reduced mobility of the ossicles:** The ossicles are the three small bones in the middle ear that transmit sound waves to the inner ear. Reduced mobility of the ossicles can be caused by infection, otosclerosis, or other conditions.<br>• **Middle ear effusion:** This is a collection of fluid in the middle ear. It can be caused by infection, allergies, or other conditions.<br>The treatment for conductive hearing loss depends on the underlying cause. If the hearing loss is caused by a simple problem, such as earwax buildup, it can be treated with a simple procedure to remove the earwax. However, if the hearing loss is caused by a more serious condition, such as an infection or otosclerosis, surgery may be necessary.<br>In some cases, hearing aids can also be used to improve hearing in people with conductive hearing loss. However, hearing aids are not always effective in treating conductive hearing loss, and they may not be able to completely restore hearing.<br>If you are diagnosed with conductive hearing loss, it is important to work with your doctor or audiologist to develop a treatment plan that is right for you. |
| 8 | Does it make sense to do a auditory brainstem responses | Yes, it does make sense to do an auditory brainstem response (ABR) examination if an MRI confirmed a large cerebellopontine angle tumor in one ear. ABR is a test that measures the electrical activity of the brain in response to sound. It can be used to assess the function of the auditory nerve and the brainstem, which are the parts of the hearing system that can be affected by a tumor in the cerebellopontine angle. |

| | | |
|---|---|---|
| | examination if an MRI confirmed large cerebello pontine angle tumor in one ear? | In a person with a normal hearing system, the ABR waves are well-defined and occur at specific times after the sound stimulus is presented. However, in people with hearing loss caused by a tumor in the cerebellopontine angle, the ABR waves may be absent or abnormal. This can help to confirm the diagnosis of hearing loss caused by a tumor and to assess the severity of the hearing loss.<br>In addition, ABR can be used to monitor the progression of hearing loss in people with a tumor in the cerebellopontine angle. If the ABR waves become more abnormal over time, it may indicate that the tumor is growing and is affecting the function of the auditory nerve and the brainstem.<br>Therefore, ABR is a valuable test that can be used to assess hearing loss caused by a tumor in the cerebellopontine angle. It can also be used to monitor the progression of hearing loss and to help guide treatment decisions.<br>Here are some of the benefits of ABR testing in people with cerebellopontine angle tumors:<br>• It can help to confirm the diagnosis of hearing loss caused by a tumor.<br>• It can assess the severity of the hearing loss.<br>• It can be used to monitor the progression of hearing loss.<br>• It can help to guide treatment decisions.<br>If you have a cerebellopontine angle tumor and you are experiencing hearing loss, you should talk to your doctor about ABR testing. They can help you determine if ABR testing is right for you and can explain the benefits and risks of the test. |
| 9 | What is the therapeutic management of acoustic neuroma? | Acoustic neuroma, also known as vestibular schwannoma, is a slow-growing, non-cancerous tumor that arises from the Schwann cells of the vestibulocochlear nerve (CN VIII). It is the most common tumor of the cerebellopontine angle (CPA), the area where the brainstem meets the cerebellum.<br>The therapeutic management of acoustic neuroma depends on a number of factors, including the size of the tumor, the patient's age and overall health, and the patient's preferences.<br>• **Observation:** If the tumor is small and not causing any symptoms, it may be monitored with regular MRI scans. This is called active surveillance or watchful waiting.<br>• **Surgical removal:** If the tumor is growing or causing symptoms, it may be removed surgically. Surgery is the most effective way to remove the tumor completely. However, it can also damage the facial nerve and hearing nerve, which can lead to hearing loss and facial weakness.<br>• **Stereotactic radiosurgery:** Stereotactic radiosurgery is a type of radiation therapy that uses high-dose radiation to destroy the tumor cells. It is a less invasive option than surgery, but it may not be as effective in completely removing the tumor.<br>The decision of whether to observe, operate, or use radiosurgery is a complex one that should be made after careful consideration of all the factors involved. The patient's age, overall health, and preferences should also be taken into account.<br>Here are some of the benefits and risks of each treatment option:<br>• **Observation:** The benefits of observation include that it is a minimally invasive option with no risk of surgery or radiation. The risks of observation include that the tumor may continue to grow and cause symptoms, and that it may become inoperable.<br>• **Surgery:** The benefits of surgery include that it is the most effective way to remove the tumor completely. The risks of surgery include damage to the facial nerve and hearing nerve, which can lead to hearing loss and facial weakness.<br>• **Stereotactic radiosurgery:** The benefits of stereotactic radiosurgery include that it is a less invasive option than surgery with a lower risk of complications. The risks of stereotactic radiosurgery include that the tumor may not be completely destroyed, and that it may cause hearing loss and facial weakness. |

| | | |
|---|---|---|
| | | If you have been diagnosed with an acoustic neuroma, you should talk to your doctor about the best treatment option for you. They can help you understand the benefits and risks of each option and make the decision that is right for you. |
| 10 | What is hidden Hearing loss? | Hidden hearing loss, also known as "auditory neuropathy spectrum disorder" (ANSD), is a type of hearing loss that is not detected by standard hearing tests. People with hidden hearing loss may have normal hearing thresholds, but they may have difficulty understanding speech in noisy environments.<br>Hidden hearing loss is caused by damage to the auditory nerve, which is responsible for carrying sound signals from the inner ear to the brain. The damage to the auditory nerve can be caused by a variety of factors, including:<br>• **Noise exposure:** Loud noise can damage the auditory nerve.<br>• **Certain medications:** Some medications can damage the auditory nerve.<br>• **Genetics:** Some people are born with a genetic condition that makes them more likely to develop hidden hearing loss.<br>• **Autoimmune diseases:** Some autoimmune diseases, such as multiple sclerosis, can damage the auditory nerve.<br>The symptoms of hidden hearing loss can vary depending on the severity of the damage to the auditory nerve. Some common symptoms include:<br>• Difficulty understanding speech in noisy environments<br>• Difficulty following conversations<br>• Difficulty hearing high-pitched sounds<br>• Fatigue<br>• Dizziness<br>• Headache<br>Hidden hearing loss can be difficult to diagnose because standard hearing tests may not detect it. However, there are a number of tests that can be used to diagnose hidden hearing loss, including:<br>• **Otoacoustic emissions (OAEs):** OAEs are sounds that are produced by the inner ear. In people with hidden hearing loss, the OAEs may be absent or abnormal.<br>• **Auditory brainstem response (ABR):** The ABR is a test that measures the electrical activity of the brain in response to sound. In people with hidden hearing loss, the ABR waves may be absent or abnormal.<br>• **Magnetic resonance imaging (MRI):** MRI can be used to look for damage to the auditory nerve.<br>There is no cure for hidden hearing loss. However, there are a number of treatments that can help to improve hearing and speech understanding, including:<br>• **Hearing aids:** Hearing aids can amplify sound, making it easier to hear.<br>• **Speech therapy:** Speech therapy can help people with hidden hearing loss to improve their listening and communication skills.<br>• **Cochlear implants:** Cochlear implants are electronic devices that can be surgically implanted in the inner ear. They can help people with severe hearing loss to hear.<br>If you think you may have hidden hearing loss, you should talk to your doctor or an audiologist. They can help you diagnose the condition and recommend the best treatment option for you. |
