## Supplementary File 4 for "Comparison of the audiological knowledge of three chatbots – ChatGPT, Bing Chat, and Bard"

Supplementary Table 1. Scores given to chatbot responses.

| question no. | ChatGPT | BingChat | Bard |
|---|---|---|---|
| 1 | 4 | 4 | 3 |
| 2 | 3 | 4 | 4 |
| 3 | 4 | 3 | 1 |
| 4 | 5 | 3 | 3 |
| 5 | 5 | 4 | 4 |
| 6 | 5 | 5 | 4 |
| 7 | 5 | 4 | 5 |
| 8 | 4 | 3 | 5 |
| 9 | 5 | 4 | 5 |
| 10 | 5 | 3 | 2 |
| sum | 45 | 37 | 36 |

Supplementary Table 2. Number of detected errors in each response.

| question no. | ChatGPT | BingChat | Bard |
|---|---|---|---|
| 1 | 0 | 0 | 1 |
| 2 | 1 | 0 | 0 |
| 3 | 0 | 1 | 0 |
| 4 | 0 | 0 | 0 |
| 5 | 0 | 0 | 0 |
| 6 | 0 | 0 | 0 |
| 7 | 0 | 0 | 0 |
| 8 | 1 | 0 | 0 |
| 9 | 0 | 0 | 0 |
| 10 | 0 | 0 | 5 |
| sum | 2 | 1 | 6 |

Supplementary Table 3. Number of detected inaccuracies in each response.

| question no. | ChatGPT | BingChat | Bard |
|---|---|---|---|
| 1 | 1 | 1 | 1 |
| 2 | 0 | 0 | 0 |
| 3 | 2 | 1 | 0 |
| 4 | 0 | 1 | 1 |
| 5 | 0 | 0 | 0 |
| 6 | 0 | 0 | 0 |
| 7 | 0 | 0 | 0 |
| 8 | 1 | 0 | 0 |
| 9 | 0 | 0 | 0 |
| 10 | 0 | 1 | 0 |
| sum | 4 | 4 | 2 |

Supplementary Table 4. Does the response cited the source (yes – 1, no – 0).

| question no. | ChatGPT | BingChat | Bard |
|---|---|---|---|
| 1 | 0 | 1 | 1 |
| 2 | 0 | 1 | 1 |
| 3 | 0 | 1 | 0 |
| 4 | 0 | 0 | 0 |
| 5 | 0 | 1 | 0 |
| 6 | 0 | 1 | 0 |
| 7 | 0 | 1 | 0 |
| 8 | 0 | 1 | 0 |
| 9 | 0 | 0 | 0 |
| 10 | 0 | 1 | 0 |
| sum | 0 | 8 | 2 |

Supplementary Table 5. Does the response suggested consulting a specialist (yes – 1, no – 0).

| question no. | ChatGPT | BingChat | Bard |
|---|---|---|---|
| 1 | 0 | 0 | 1 |
| 2 | 0 | 0 | 1 |
| 3 | 0 | 0 | 0 |
| 4 | 0 | 0 | 1 |
| 5 | 1 | 0 | 1 |
| 6 | 0 | 0 | 1 |
| 7 | 1 | 0 | 1 |
| 8 | 1 | 1 | 1 |
| 9 | 0 | 0 | 1 |
| 10 | 1 | 0 | 1 |
| sum | 4 | 1 | 9 |

Supplementary Table 6. Number of words in a response.

| question no. | ChatGPT | BingChat | Bard |
|---|---|---|---|
| 1 | 534 | 160 | 358 |
| 2 | 464 | 107 | 382 |
| 3 | 418 | 175 | - |
| 4 | 494 | 69 | 366 |
| 5 | 515 | 169 | 355 |
| 6 | 388 | 145 | 255 |
| 7 | 482 | 72 | 383 |
| 8 | 450 | 86 | 335 |
| 9 | 535 | 54 | 405 |
| 10 | 443 | 88 | 399 |
| mean | 472.3 | 112.5 | 359.8 |
| std | 49.0 | 45.6 | 45.1 |
